# Machine learning-based imaging biomarkers improve statistical power in clinical trials

**DOI:** 10.1101/19010041

**Authors:** Carolyn Lou, Mohamad Habes, Christos Davatzikos, Russell T. Shinohara, Alzheimer’s Disease Neuroimaging Initiative

## Abstract

Radiomic models, which leverage complex imaging patterns and machine learning, are increasingly accurate in predicting patient response to treatment and clinical outcome on an individual patient basis. In this work, we show that this predictive power can be utilized in clinical trials to significantly increase statistical power to detect treatment effects or reduce the sample size required to achieve a given power. Akin to the historical control paradigm, we propose to utilize a radiomic prediction model to generate a pseudo-control sample for each individual in the trial of interest. We then incorporate these pseudo-controls into the analysis of the clinical trial of interest using classical and well established statistical tools, and investigate statistical power. Effectively, this approach utilizes each individual’s radiomics-based predictor of outcome for comparison with the actual outcome, potentially increasing statistical power considerably, depending on the accuracy of the predictor. In simulations of treatment effects based on real radiomic predictive models from brain cancer and prodromal Alzheimer’s Disease, we show that this methodology can decrease the required sample sizes by as much as a half, depending on the strength of the radiomic predictor. We further find that this method is most helpful when treatment effect sizes are small and that power grows with the accuracy of radiomic prediction.

---

**I**n recent decades, rapid advances in technology have increased the amount of neuroimaging data available to researchers at an unprecedented rate (1, 2). Machine learning methods empower the integration of this high-dimensional data into powerful individualized predictive markers that have been shown to be useful for tasks such as diagnosis and prognosis in diseases such as Alzheimer’s disease and brain cancers (3, 4). Predictive modeling is poised to receive the benefits of the large and varied nature of this data.

With the growing availability of big data in medical imaging, a central focus has emerged on the development of increasingly complex tools for their analysis with the primary goal of individualized predictions (5). In this paper, we propose harnessing these powerful machine learning tools for the analysis of clinical trials by using them as a means to inform statistical analyses with individualized estimates of clinical outcome. We therefore arrive at the concept of individualized evaluation of treatment effects in clinical trials.

There is an extensive literature on the use of historical controls to supplement data from new clinical trials that have largely relied on pooling methods or Bayesian modeling (6, 7). Whereas these methods augment data for a trial by incorporating historical data on the group level, high-dimensional predictors offer the opportunity to augment current trials by incorporating historical data in the form of individualized predictions at the individual level. This allows for a more precise evaluation of the treatment effect for each person, rather than relying on a group-level effect that determines average outcome.

Here, we present a method that draws on these ideas while leveraging powerful predictive biomarkers and the wealth of data used to build them to generate personalized predictions of outcome. These predictions can be used directly in the analysis of data in clinical trials. We find that this methodology can substantially improve statistical power for detecting treatment effects, depending on the predictive power of the machine learning-based model. Correspondingly, this approach can substantially reduce the sample size needed to achieve the same power in a clinical trial.

## Methods

Our method relies on access to two sets of data: i) a current clinical trial designed to study an outcome of interest and ii) a cohort of similar subjects treated according to the current standard of care. We narrow our focus in this work to radiomic predictors and associated studies, so we assume that imaging data has been gathered at study enrollment for both sets of trials. However, more broadly we only require a predictive model that is based on sufficient information measured at baseline on each participant in both datasets to predict the outcome under standard of care. The techniques proposed here are also directly applicable to other -omic modeling scenarios, and generally, to any predictive marker of standard of care outcome.

Our basic premise herein is that we can utilize previously collected imaging data to build a radiomic prediction model, fully validate it, and use it to generate a single score that summarizes imaging patterns that predict future clinical outcome of interest, such as patient survival, progression-free survival, or response to treatment (Figure 1). The model that is built based on the historical trial can then be used in conjunction with data collected from the current trial to generate individualized values of the radiomic score for each of the current participants. These individualized scores represent predicted values for how the treated individuals in the current trial would have fared had they instead been assigned to the control group. The incorporation of these predicted values lends power to the detection of the effect of a treatment in the final analysis of the current trial by modeling the inter-subject variability in the outcome in terms of baseline heterogeneity represented in the baseline imaging.

**Figure 1.**
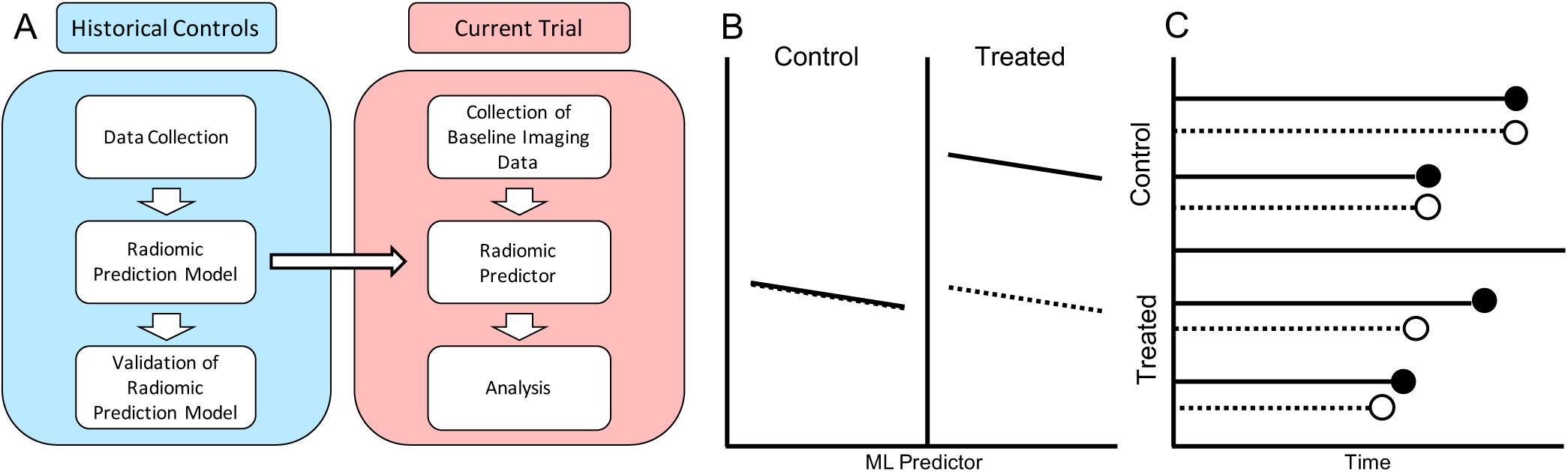
A: Workflow for implementing the proposed method in a new clinical trial. B (continuous) and C (survival outcome): Schematic diagram for individualized predictions that are generated for each person in the current trial, where the solid lines indicate observed outcome for the participants of the current trial and the dashed lines indicate predicted outcome for those participants had they not been treated.

To investigate the advantage of this approach, we consider two scenarios. To more closely approximate real-life clinical trial performance, we use radiomic and outcome data from two observational studies to generate hypothetical study data, where the first focuses on continuous outcomes of cognitive decline in prodromal Alzheimer’s disease (AD) and the second on survival after diagnosis with glioblastoma multiforme (GBM). In these studies, we randomly split the data into a historical cohort and a trial cohort and then simulate effects in a randomly selected subset (corresponding to one arm) of the trial cohort. We then compare the statistical power of our proposed approach with the classical modeling approach that does not include radiomic prediction-based modeling.

### Data

In our first study, we consider the case of therapeutic trials for AD in which the outcome is longitudinal cognitive change. We utilize data derived from the Azheimer’s Disease Neuroimaging Initiative (ADNI, adni.loni.usc.edu) on 400 subjects with mild cognitive impairment (MCI) who underwent serial MRI at 1.5T ^∗^. Data used from this study consisted of cross-validated predictions of time to AD diagnosis using the SPARE-AD score (8). SPARE-AD is derived from patterns of regional brain atrophy (volume loss) captured by atlas warping methods and high-dimensional pattern classification using support vector machines (SVM) aiming to differentiate cognitively normal and Alzheimer’s disease subjects (3, 9). The outcome of interest here is cognitive decline as measured by 3-year change from baseline values of the ADNI composite memory score (10) (ADNI-MEM). Of the 400 MCI subjects in our study, 283 have 3-year ADNI-MEM scores available. Average 3-year change from baseline for ADNI-MEM in the current study was -0.17 (standard deviation 0.49).

As a second case study, we focus on therapeutic trials for GBM therapies, which aim to prolong survival after diagnosis. We conduct our simulated treatment study using 134 patients who were treated for newly diagnosed GBM at the Hospital of the University of Pennsylvania between 2006 and 2013. The actual median survival in this sample was 12 months, and survival data were assessed for all subjects with no loss to follow-up. Detailed demographics and a clinical description of these subjects have been previously published (4). For studies involving these data, we investigate the use of cross-validated predictions of survival time based on radiomic analyses of pre- and post-contrast T1-weighted, FLAIR, diffusion, and perfusion imaging acquired pre-operatively at diagnosis. This GBM predictive model utilizes an SVM to differentiate short, medium, and long survival (4).

### Statistical Methods

All hypothesis testing is conducted assuming a 5% type I error rate and using two-sided alternatives. For our continuous outcome analyses, we apply linear regression modeling of the outcome and employ Wald tests to assess whether treatment groups differed in their outcomes either i) adjusting for the radiomic predictor by inclusion as covariate, or the classical approach with corresponds to ii) not adjusting for the radiomic predictor. For time-to-event outcomes, we assess differences between treatment groups with and without adjustment for the radiomic prediction by assuming an accelerated failure time model.

We conduct two sets of real data simulations: one set focusing on cognitive decline in AD, and one set focusing on GBM survival outcomes. For both, we sample without replacement twice from the observed data: for the first group indexed by *i* = 1, …, *n/*2, we set our treatment indicator *A*_*i*_ = 0 and record the observed outcome *Y*_*i*0_, as well as the value of the radiomic predictor *X*_*i*_ at baseline. For the second group, indexed by *i* = *n/*2 +1, …, *n*, we introduce a treatment effect *γ*, set our treatment indicator *A*_*i*_ = 1, and again record outcome *Y*_*i*0_ and baseline radiomic predictor measurement *X*_*i*_. We repeat this process 1000 times, recording the p-value corresponding to the test for treatment effect each time. We calculate type I error rate and power as the percentage of time the treatment effect is significant at the *α* = 0.05 level, where *γ* is set to 0 to assess type I error and a non-zero value to assess power. In order to quantify the sample size benefits from using this method, we repeat the above procedure for a range of sample sizes *n*, and the smallest *n* for which power reaches 80% is recorded. We explore this for a range of hypothetical effect sizes, which is defined here as *γ* divided by the standard deviation of the outcome.

## Results

For both continuous and time-to-event outcomes, we find that the proposed method reduces the minimum sample size *n* required for 80% power in clinical trial analyses (Figure 2). The inclusion of the imaging biomarker tends to be most helpful in terms of the absolute differences when the effect sizes are small. Type I error remains controlled throughout all experiments conducted. In the ADNI study, the classical analysis requires 16% to 18% more samples than our proposed method. In the GBM study, the classical analysis requires around 73% to 94% more samples than our proposed method.

**Figure 2.**
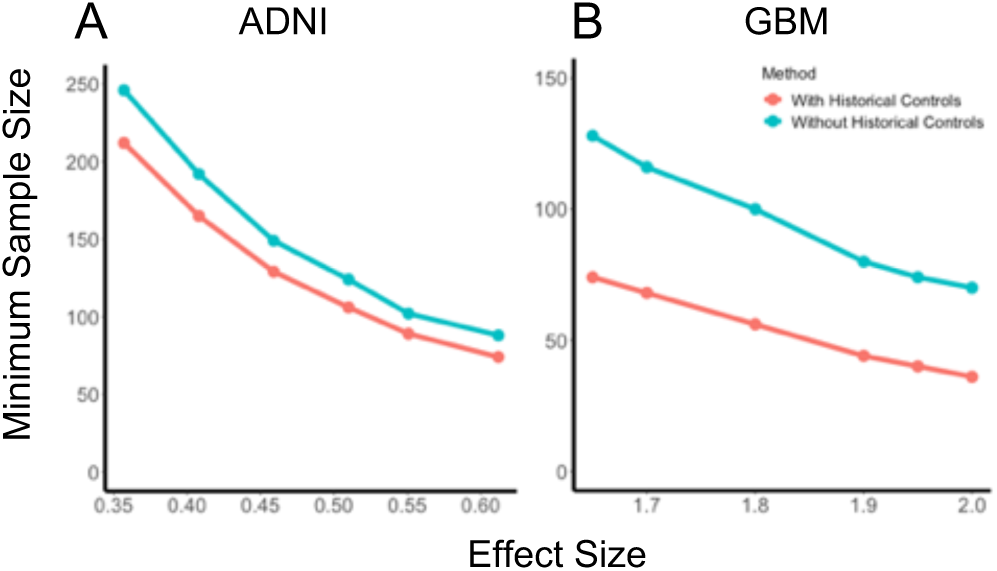
Results from simulated studies under two scenarios. With the addition of historical controls, the required sample size for 80% power is markedly lower than using classical two-sample clinical trial analysis.

## Discussion

We have shown that individualized machine learning-based imaging biomarkers can be a useful tool in a clinical trial analysis, offering an increase in power and/or a reduction of the required sample size. The novelty of this method arises from the incorporation of individualized predictions, which derive their usefulness from the powerful predictive algorithms they are based upon. As neuroimaging biomarkers derived via machine learning become more common, the set of historical data for which we have biomarker values also becomes larger, which promises to strengthen the radiomic prediction model that we use to generate predicted values.

Here, we used two previously developed biomarkers, one of which was trained to classify an outcome different from the target of the clinical trial analysis, and another which was trained to classify the same outcome as the clinical trial analysis. While both predictors offered gains in sample size reduction, predictors built specifically for the outcome of interest in the clinical trial are likely to perform better and offer more substantial gains.

The approach proposed in this paper does have some limitations. First, the use of radiomic predictions can be hindered by the cost of collecting imaging data (11). Furthermore, insufficient performance of the radiomic prediction can result in more modest improvements in (or, in extreme cases, even loss of) statistical power. Cost-benefit analyses are thus warranted. Finally, if a radiomic predictor is trained on data from a different population compared with those studied in the current trial, the improvements in statistical power may be less pronounced. However, due to the randomization in the study, the type I error rate is expected to be maintained and internal validation or calibration of the predictive model is possible using data from the control arm of a clinical trial.

Further studies of the misspecification of the predictive model as well as the clinical trial outcome model are warranted for assessing potential gains and loss of power in these settings. However, misspecification of clinical outcome models can be guarded against using statistical models satisfying symmetry criteria (12). The biomarkers from the two cases presented here were both built using SVMs, but this methodology can accommodate predictions more generally. Incorporation of these biomarkers into a one-arm trial designs in which all participants in the trial are treated similarly also requires further statistical research.

## Data Availability

All SPARE-AD scores used herein have been uploaded to http://adni.loni.ucla.edu/. All image processing software used to derive SPARE-AD, importantly the DRAMMS deformable registration and COMPARE classification pipelines, are freely available for download under http://www.rad.upenn.edu/sbia, and involve fully automated procedures.
Data for the GBM analysis are not publicly available.

http://www.rad.upenn.edu/sbia

## ACKNOWLEDGMENTS

This work was supported in part by NIH grants R01NS085211, R01MH112847, R01NS060910, RF1AG054409, R01EB022573, R01NS042645, and R01MH112070.

## Footnotes

∗ The ADNI was launched in 2003 as a public-private partnership, led by Principal Investigator Michael W. Weiner, MD. For up-to-date information, see www.adni-info.org.

